# Rare disease gene association discovery from burden analysis of the 100,000 Genomes Project data

**DOI:** 10.1101/2023.12.20.23300294

**Authors:** Valentina Cipriani, Letizia Vestito, Emma F Magavern, Julius OB Jacobsen, Gavin Arno, Elijah R Behr, Katherine A Benson, Marta Bertoli, Detlef Bockenhauer, Michael R Bowl, Kate Burley, Li F Chan, Patrick Chinnery, Peter Conlon, Marcos Costa, Alice E Davidson, Sally J Dawson, Elhussein Elhassan, Sarah E Flanagan, Marta Futema, Daniel P Gale, Sonia García-Ruiz, Cecilia Gonzalez Corcia, Helen R Griffin, Sophie Hambleton, Amy R Hicks, Henry Houlden, Richard S Houlston, Sarah A Howles, Robert Kleta, Iris Lekkerkerker, Siying Lin, Petra Liskova, Hannah Mitchison, Heba Morsy, Andrew D Mumford, William G Newman, Ruxandra Neatu, Edel A O’Toole, Albert CM Ong, Alistair T Pagnamenta, Shamima Rahman, Neil Rajan, Peter N Robinson, Mina Ryten, Omid Sadeghi-Alavijeh, John A Sayer, Claire L Shovlin, Jenny C Taylor, Omri Teltsh, Ian Tomlinson, Arianna Tucci, Clare Turnbull, Albertien M van Eerde, James S Ware, Laura M Watts, Andrew R Webster, Sarah K Westbury, Sean L Zheng, Mark Caulfield, Damian Smedley

## Abstract

To discover rare disease-gene associations, we developed a gene burden analytical framework and applied it to rare, protein-coding variants from whole genome sequencing of 35,008 cases with rare diseases and their family members recruited to the 100,000 Genomes Project (100KGP). Following *in silico* triaging of the results, 88 novel associations were identified including 38 with existing experimental evidence. We have published the confirmation of one of these associations, hereditary ataxia with *UCHL1*, and independent confirmatory evidence has recently been published for four more. We highlight a further seven compelling associations: hypertrophic cardiomyopathy with *DYSF* and *SLC4A3* where both genes show high/specific heart expression and existing associations to skeletal dystrophies or short QT syndrome respectively; monogenic diabetes with *UNC13A* with a known role in the regulation of β cells and a mouse model with impaired glucose tolerance; epilepsy with *KCNQ1* where a mouse model shows seizures and the existing long QT syndrome association may be linked; early onset Parkinson’s disease with *RYR1* with existing links to tremor pathophysiology and a mouse model with neurological phenotypes; anterior segment ocular abnormalities associated with *POMK* showing expression in corneal cells and with a zebrafish model with developmental ocular abnormalities; and cystic kidney disease with *COL4A3* showing high renal expression and prior evidence for a digenic or modifying role in renal disease. Confirmation of all 88 associations would lead to potential diagnoses in 456 molecularly undiagnosed cases within the 100KGP, as well as other rare disease patients worldwide, highlighting the clinical impact of a large-scale statistical approach to rare disease gene discovery.

---

Rare diseases collectively affect one in seventeen individuals in the United Kingdom ^1^. Despite advances in genomic sequencing, molecular diagnosis continues to elude 50% to 80% of patients presenting to genetics clinics ^2^. Furthermore, less than half of the 10,000 rare Mendelian diseases in the Online Mendelian Inheritance in Man (OMIM) database ^3^ have an established genetic basis. Diagnostic failure may arise due to a lack of routine screening for non-coding ^1^ or structural variants ^4^. However, it is likely that a substantial proportion of the pathogenic variants responsible for undiagnosed cases reside in those yet to be discovered (possibly very rare) disease-associated genes. The scale of rare disease sequencing studies such as the Undiagnosed Disease Network ^5^, Centers for Mendelian Genomics ^6^, Deciphering Developmental Disorders ^7^ and the 100,000 Genomes Project (100KGP) ^8^, offers expanded opportunities to provide insight into pathogenic mechanisms of inherited disease, including the possibility of establishing disease-gene associations through case-control analyses, akin to methods previously used to identify common genetic variants influencing the risk of complex disorders. Such an approach provides much-needed power to identify genes harbouring rare pathogenic variants.

To identify disease-associated genes, we recently developed a framework that analyses rare protein coding variants identified by the Exomiser prioritisation tool ^9^, within a preliminary version of the 100KGP data ^4^, to conduct gene-based burden testing of single probands and family members relative to control families. *In silico* triage highlighted 22 novel disease-gene associations, three of which have been also reported in independent studies ^10–12^.

We have now extended our gene burden analytical framework for generic application to any large-scale, rare disease sequencing cohorts and complemented it with visualisation scripts. In addition, we report on the application of the approach to a larger cohort from the final 100KGP data including 35,008 families, 226 rare diseases and a pool of 4,676,866 rare candidate variants with improved *in silico* and added clinical expert triage of 88 probable new disease-gene associations.

## Results

### Gene burden analytical framework for disease gene discovery

We have developed an open-source R framework (https://github.com/whri-phenogenomics/geneBurdenRD) allowing users to perform gene burden testing of variants in user-defined cases versus controls from rare disease sequencing cohorts. The input to the framework is a file obtained from processing Exomiser output files for each of the cohort samples, a file containing a label for each case-control association analysis to perform within the cohort and a (set of) corresponding file(s) with user-defined identifiers and case/control assignment per each sample. Cases and controls in a cohort could be defined in many ways, for example, by recruited disease category as we have done for the 100KGP analysis below, by specific phenotypic annotations or phenotypic clustering. The framework will then assess false discovery rate (FDR)-adjusted disease-gene associations where genes are tested for an enrichment in cases vs controls of rare, protein-coding, segregating variants that are either (i) predicted loss-of-function (LoF), (ii) highly predicted pathogenic (Exomiser variant score >= 0.8), (iii) highly predicted pathogenic and present in a constrained coding region (CCR; ^13^) or (iv) *de novo* (restricted to only trios or larger families where *de novo* calling was possible and provided by the user) (**Methods**). As well as various output files annotating these case-control association tests, Manhattan and volcano plots are generated summarising the FDR-adjusted p-values of all the gene-based tests for each case-control association analysis, along with lollipop plots of the relevant variants in cases and controls and plots of the hierarchical distribution of the Human Phenotype Ontology (HPO) case annotations for individual disease-gene associations.

### Application to the 100,000 Genomes Project data identifies 88 novel disease-gene associations

A rare variant gene-burden analysis was performed on a cohort of 35,008 families (73,018 genomes) from the 100KGP rare disease pilot and main programme (Data Release v.11) (**Fig. 1, Supplementary Table 1**). A pool of 4,676,866 rare, protein-coding, segregating and most predicted pathogenic (per gene) variants for the analysis was derived by running Exomiser for each family. Our pipeline was then used to detect statistically significant gene-based enrichment in relevant variant categories (predicted LoF, highly predicted pathogenic, highly predicted pathogenic in CCR regions, *de novo*) for *cases* in each of 226 ‘specific diseases’ used for patient recruitment by the 100KGP ^4^ versus *controls* who were defined as probands from any other 20 broad ‘disease groups’ in the project (e.g. intellectual disability cases were compared to all non-neurological probands as controls). Applied cutoffs required at least 5 case probands per ‘specific disease’ and at least 4 probands (of which, at least one *case*) with a relevant rare variant per disease-gene burden test (**Methods**).

**Figure 1.**
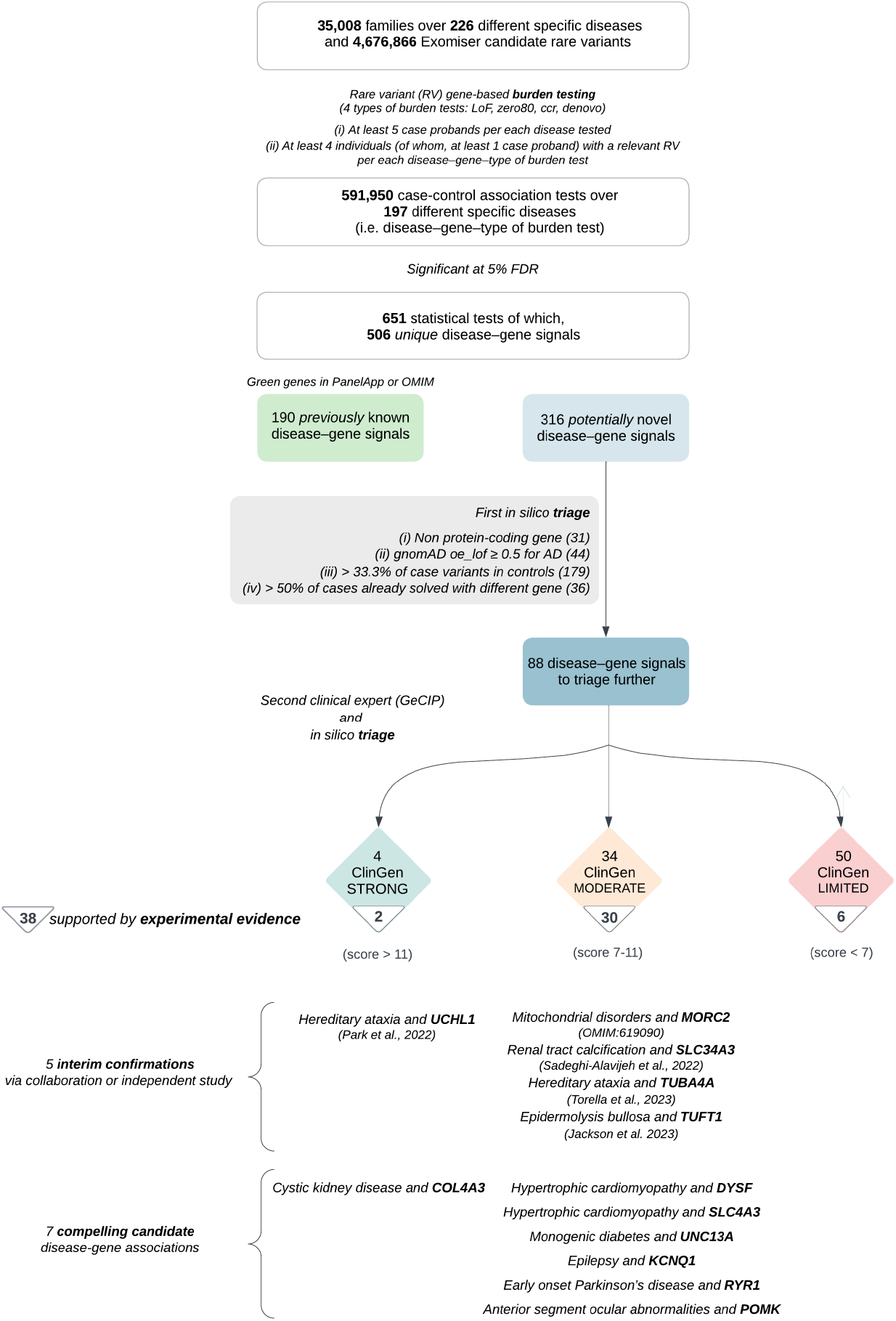
Rare variant gene burden analysis of the 100,000 Genomes Project data. Flowchart of the rare variant gene-based analytical framework, including triaging of the results.

We identified 190 previously known and 316 novel potential disease-gene associations (**Fig. 2**), imposing a 5% FDR. Not previously known (novel) signals were initially defined, at the first round of analysis in March 2021, as having no documented evidence for an association within OMIM and absence from the ‘specific disease’ curated panel of high confidence (green) genes in PanelApp ^14^. Enrichment of predicted LoF, highly predicted pathogenic, also in CCR and *de novo* variants was observed in 53%, 28%, 6% and 13% of known and 19%, 80%, 1%, and 0% of novel associations respectively, revealing discovery was mostly driven by predicted pathogenic, missense variants.

**Figure 2.**
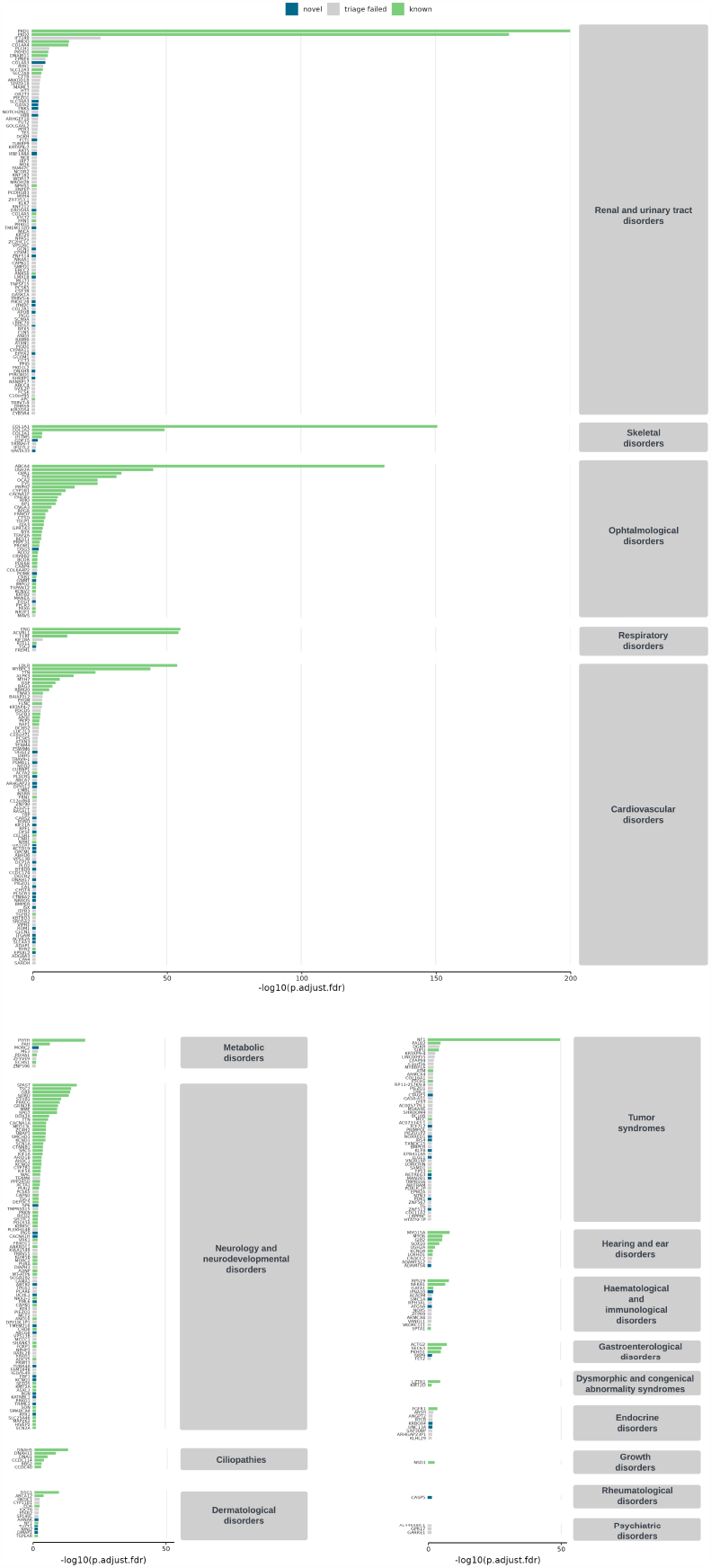
Rare disease gene discoveries from gene burden analysis of the 100,000 Genomes Project data. The gene burden testing identified 506 disease-gene associations at 5% False Discovery Rate (FDR) including 316 potentially novel. An initial triage of the novel signals identified 88 signals for further investigation through *in silico* collection for additional evidence and clinical expert review. Statistical significance, expressed as -log10 of FDR adjusted p-value, is shown for each of the 506 gene-disease associations significant at 5% FDR, arranged by ‘disease group’. The 190 known associations are in green, 88 triaged novel signals in blue and the discarded signals in grey.

Potential associations were further filtered to 88/316 (28%) by removing those in which the gene was a non-protein coding RNA one (31/316); (ii) for signals driven by dominant, LoF variants where the Genome Aggregation Database (gnomAD) ^15^ suggests there is no evidence for haploinsufficiency (i.e. gnomAD oe_lof >= 0.5) (44/316); (iii) > 33.3% of the variants driving the signal in the cases were present in controls (179/316) to exclude false positives unless penetrance is very low; or (iv) > 50% of the cases driving the signals had already received an alternative genetic diagnosis (36/316) (**Fig. 1** and **Supplementary Table 2**). Molecularly diagnosed individuals with a report as part of the 100KGP routine diagnostic pipeline were included in the experimental pipeline to evaluate the approach via detection of known associations and to allow for the possibility of misdiagnosis. Relatively relaxed thresholds were chosen for the last two criteria (iii and iv) to avoid restricting associations based on two or three cases, but most were far below the relevant thresholds, e.g. 48/88 had no occurrence of putative disease-causing variants in controls and in 50/87 less than 5% of cases were otherwise assigned a molecular diagnosis. However, for all 12 cystic kidney disease associations 20-50% of cases had received a diagnosis involving well known genes such as *PKD1*, raising the possibility of digenic inheritance or variants in other genes modifying penetrance as a common theme (discussed further for the *COL4A3* example below). Similarly, most of the independent Irish renal cohort cases described in **Supplementary Table 2** already had an established molecular diagnosis. In comparison to the novel association signals, 174/190 (92%) of signals from known disease genes passed criteria from (i) to (iii). Given the relatively mixed ethnic diversity of the UK population and our cohort, we investigated if any of the 88 associations in **Supplementary Table 2** were linked to particular ancestries. For most, no significant enrichments were detected, but the *DYSF* association had 25% Asian/Asian British and 12.5% Black/Black British cases, the *DSG3* association had 27% Black/Black British cases, and both *IFN10* associated cases were Black/Black British. Different variants were observed in all these examples.

An extensive review of the literature as well as the phenotype evidence from Exomiser, in collaboration with members of the Genomics England Clinical Interpretation Partnerships (GeCIPs), was performed to identify supporting evidence from each of the 88 genes’ biological function, known disease associations or the phenotypes of the gene-deficient mouse and/or other animal models. *In silico* analyses were also undertaken to identify high quality StringDB ^16^ protein-protein associations between the gene signal and any other genes known to be associated with the disease, or with highly specific expression in the most relevant tissue for the disease. This combined curation highlighted 38 associations supported by experimental evidence: 29 based on literature curation of a gene function fitting the likely disease mechanism with additional lines of evidence for many, seven based on mouse models and other evidence for some, and two based on protein-protein evidence only (**Fig. 1** and highlighted in bold in the summary column in **Supplementary Table 2**).

ClinGen ^17^ has developed a robust set of criteria to assess the evidence for disease-gene associations and we applied these to our 88 associations. Evidence of causality was strong for four associations, moderate for 34, and limited for the remainder (**Fig. 1 and Supplementary Table 2**). The four associations with strong evidence were familial thoracic aortic aneurysm disease and *UGGT2*; cystic kidney disease with *DNAH8* and *COL4A3*; hereditary ataxia and *UCHL1*. The latter two are described below but the first two are driven almost purely by the genetic evidence score due to many case variants, with little or no experimental evidence.

Four of our 88 novel signals have had interim independent confirmatory evidence emerge since our initial assessment was performed in 2021: hereditary ataxia associated with *TUBA4A* ^18^; mitochondrial disorders with *MORC2* ^19^; renal tract calcification with *SLC34A3* ^20^; and epidermolysis bullosa with *TUFT1* ^21^. In an interim published collaborative work ^22^, we were also able to confirm the association between heterozygous variants in *UCHL1* and a specific form of hereditary ataxia associated with neuropathy and optic atrophy, which is distinct from the spastic paraplegia associated with recessive variants in the same gene. The signal was driven by rare LoF variants in five cases and the gnomAD haploinsufficiency evidence is particularly compelling with no observed LoF variants (o/e = 0 (0-0.21) and pLI = 0.99). A heterozygous KO mouse model with neurological phenotypes also exists ^23^. Through a network of collaborators in the UK and Germany, we were able to identify a total of 34 cases from 18 unrelated families and confirmed a 50% reduction in expression through mass spectrometry-based proteomics on patient fibroblasts.

Of the remaining 83 novel associations, nine had prior functional data fitting the likely disease mechanism and a high ClinGen classification score (>= 8) (**Supplementary Table 2**). Further investigation of the association between congenital heart disease and *DCP1A* revealed that the case variants are likely to be false positive calls resulting from two neighbouring inframe deletions. Investigation of the recessive association between *HBB* variants and renal tract calcification highlighted that nearly all the identified variants are reported to be known pathogenic/likely pathogenic in ClinVar and linked to beta-thalassemia; the patients included in our analyses were also clinically described with thalassemia. Therefore, despite previous descriptions of the association of *HBB* with kidney stones ^24^ and nephropathy of unknown cause ^25^, we propose that our analyses have detected an association of thalassemia with renal tract calcification, appearing as a new association possibly more due to a treatment effect ^26^. The remaining seven novel compelling candidates are described in the following sections.

### Hypertrophic cardiomyopathy associated with *DYSF*

We identified a dominant association between variants in the muscle fiber repair gene Dysferlin (*DYSF)* and ‘specific disease’ *hypertrophic cardiomyopathy* (HCM). The association is driven by rare, predicted pathogenic variants throughout the gene (22 missense, 1 LoF, 1 intronic) in 24 cases (**Fig. 3a**, odds ratio (OR) = 2.8 [95% confidence interval (CI): 1.8 – 4.3], **Supplementary Table 2**). To assess if participants with predicted pathogenic mutations in this gene had a phenotypically distinct sub-endotype, we performed pairwise similarity by HPO term calculations of all the HCM cases as described in the **Methods**. The 24 cases associated with the *DYSF* signal shared a similar clinical syndrome (mean PhenoDigm score from pairwise, reciprocal, non-self hits was 0.67) with respiratory and metabolic phenotypes in addition to the cardiomyopathy. However, 301/1,000 randomly sampled HCM sets of the same size also achieved the same mean score or higher, suggesting the *DYSF* families are not a phenotypically distinct sub-group of HCM based on the available HPO annotations. Co-segregation was apparent in two trios where (i) a heterozygous, ClinVar known pathogenic intronic c.5667-824C>T variant ^27^ is inherited from the affected father in the male proband, (ii) a heterozygous p.Pro810Ser variant, classified as a variant of uncertain significance (VUS) in ClinVar, is inherited from the affected father in the male proband (**Fig. 3a**). *DYSF* shows strongest expression in skeletal muscle, but with significant levels of expression in heart muscle as well (**Fig. 3a**). In addition, co-expression network analysis of *DYSF* and known HCM genes from PanelApp shows the most significant enrichment in the heartAtrial Genotype Tissue Expression (GTEx) v6 module (FDR adjusted p-value of 3.6×10^−13^), with *DYSF* co-expressed with 10/22 of the known genes (**Fig. 3a**). LoF in *DYSF is a* known cause of autosomal recessive limb-girdle muscular dystrophy (LGMD) 2 (OMIM:253601), although there is evidence for a cardiomyopathic component and a milder, later onset form for heterozygous carriers ^28^. The association between *DYSF* and HCM driven by heterozygous missense, rather than LoF, variants suggests that a milder, cardiomyopathy-only phenotype, rather than LGMD, is possible.

**Figure 3.**
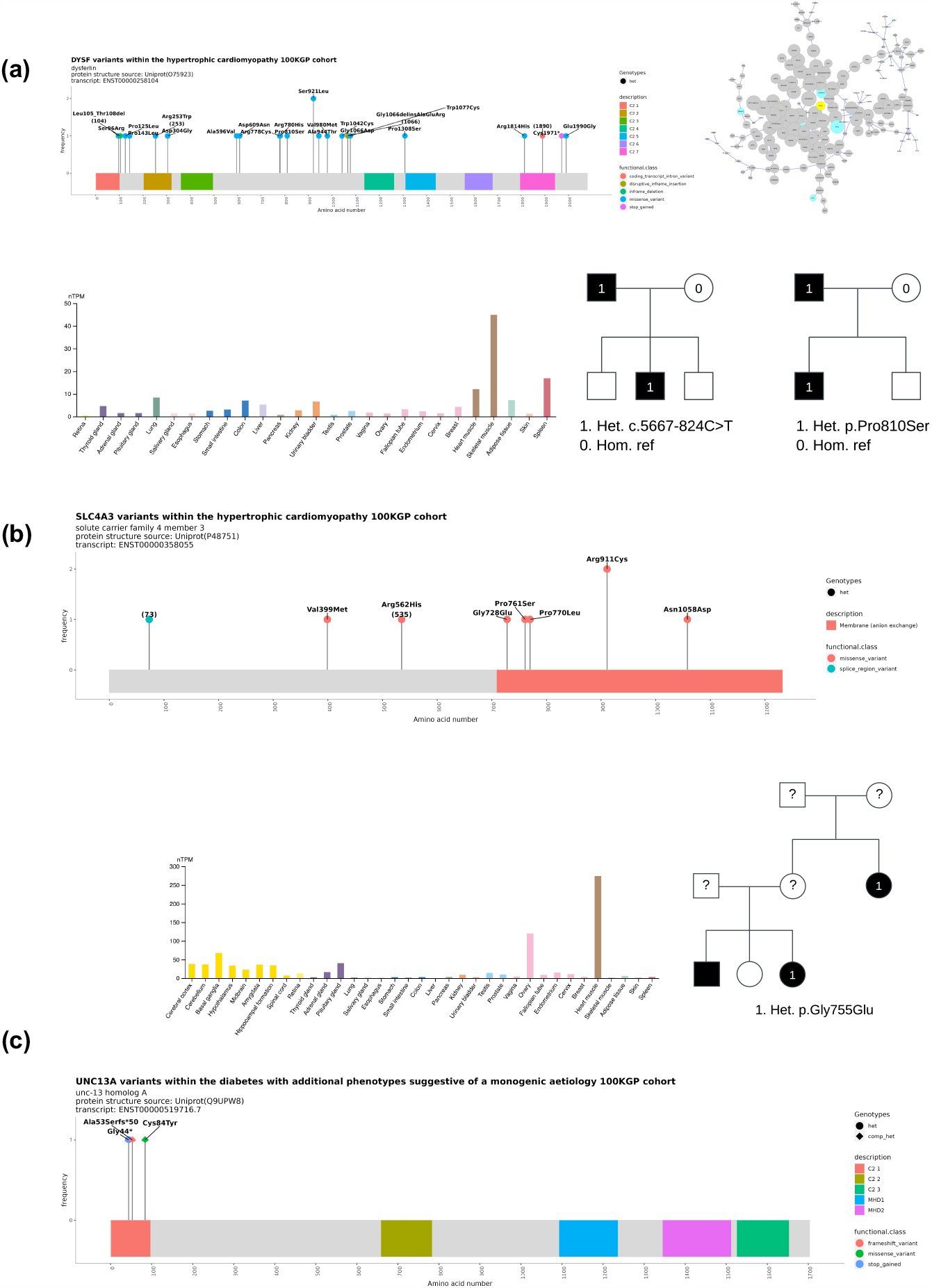
Evidence for associations based on location of rare, predicted pathogenic variants in the 100,000 Genomes Project cases, Genotype Tissue Expression (GTEx) Project data and co-segregation data. **(a)** hypertrophic cardiomyopathy with *DYSF* including the heartAtrial GTEx co-expression module with known HCM genes in blue and *DYSF* in yellow, **(b)** hypertrophic cardiomyopathy with *SLC4A3*, **(c)** monogenic diabetes with *UNC13A*.

### Hypertrophic cardiomyopathy associated with *SLC4A3*

We identified another dominant association between variants in *SLC4A3* and ‘specific disease’ HCM. The association is driven by rare, predicted pathogenic variants (8 missense, 1 splice region) in 9 cases located throughout the gene (**Fig. 3b**, OR = 5.9 [95% CI: 2.9 – 12.0], **Supplementary Table 2**). One family contains the same p.Gly728Glu variant in the proband and her affected maternal aunt. A p.Arg938His variant is seen in two of the other cases. The gene is depleted for rare missense variants in gnomAD (o/e = 0.73 (0.68-0.78) and Z = 2.74). *SLC4A3*, a plasma membrane anion exchange protein, has recently been associated with short QT syndrome ^29^ and exhibits highly specific expression in heart muscle. None of our cases had recorded short QT phenotypes, but the underlying ECG data was not available for closer evaluation. The 9 cases show strong phenotypic similarity to each other (mean PhenoDigm score from pairwise, reciprocal, non-self hits was 0.61) with palpitations/arrhythmia phenotypes observed in 6 cases in addition to the HCM. However, 596/1000 randomly sampled case sets of the same size achieved the same mean score or higher, suggesting the *SLC4A3* families are not phenotypically distinct from the other HCM cases based on the available HPO terms.

### Monogenic diabetes associated with *UNC13A*

We identified a dominant association between variants in *UNC13A* and ‘specific disease’ *diabetes with additional phenotypes suggestive of a monogenic aetiology*. The association is driven by rare predicted LoF variants in 2 singleton cases with the only recorded phenotypes being diabetes mellitus in both and one further phenotype in one: p.Ala53Serfs*50 and p.Gly44* (**Fig. 3c**, OR = 355.1 [95% CI: 71.7 – 1759.5], **Supplementary Table 2**). The gene is depleted for rare LoF variants in gnomAD (o/e = 0.09 (0.05-0.16) and pLI = 1). *UNC13A* is a diacylgycerol and phorbol ester receptor with evidence for a role in regulation of β cells ^30^. Neonatal pancreatic β cells extracted from UNC13A-knockout mice and knock-in mice that lacked the DAG binding domain showed impaired second phase of insulin secretion in response to glucose stimulation ^31^ and the heterozygous mouse knockout model shows impaired glucose tolerance ^32^. In addition, co-expression network analysis of *UNC13A* and known monogenic diabetes genes from PanelApp shows the most significant enrichment in the pancreas GTEx v6 module (FDR adjusted p-value of 0.01), with *UNC13A* co-expressed with 6/43 of the known genes. However, predicted LoF variants (3 splice site, 5 stop gain or frameshift) were also seen in controls with no apparent history of diabetes suggesting either incomplete penetrance, later onset (year of birth of the two was 1973 and 1974 compared to 1956-2007 (mean 1980) for controls), or that the variants in controls are not genuinely LoF.

### Epilepsy associated with *KCNQ1*

We identified a dominant association between variants in *KNCQ1* and ‘specific disease’ *epilepsy plus other features*. The association is driven by rare, predicted pathogenic variants (20 missense, 1 LoF) in 21 cases, located throughout the gene (**Fig. 4a**, OR = 3.0 [95% CI: 1.9 – 4.8], **Supplementary Table 2**). *KCNQ1* is depleted for rare missense variants in gnomAD (o/e = 0.82 (0.77-0.87) and Z = 1.99). The 21 cases show strong phenotypic similarity to each other (mean PhenoDigm score from pairwise, reciprocal, non-self hits was 0.63). Only 62/1000 randomly sampled epilepsy sets of the same size achieved the same mean score or higher, suggesting the *KCNQ1* families with common features of seizures, deterioration of higher mental function and cerebral morphology abnormalities are phenotypically distinct from the other epilepsy cases. The only co-segregation evidence comes from a duo where the p.His509Gln variant in the proband is inherited from an affected mother. *KNCQ1* is a voltage-gated potassium channel required for the repolarization phase of the cardiac action potential and associated with autosomal dominant long-QT syndrome sub-type 1 (OMIM:192500), while a mouse model supports the potential for associated epileptic seizures ^33^. Other voltage-gated potassium channel family members, *KCNQ2* (OMIM:613720), *KCNQ3 (*OMIM:121201) and *KCNQ5 (*OMIM:617601), have been associated with epilepsy and are on the virtual panel for this specific disease in PanelApp. However, distinguishing primary seizures from seizures secondary to cerebral hypoperfusion caused by ventricular arrhythmia is challenging ^34^.

**Figure 4.**
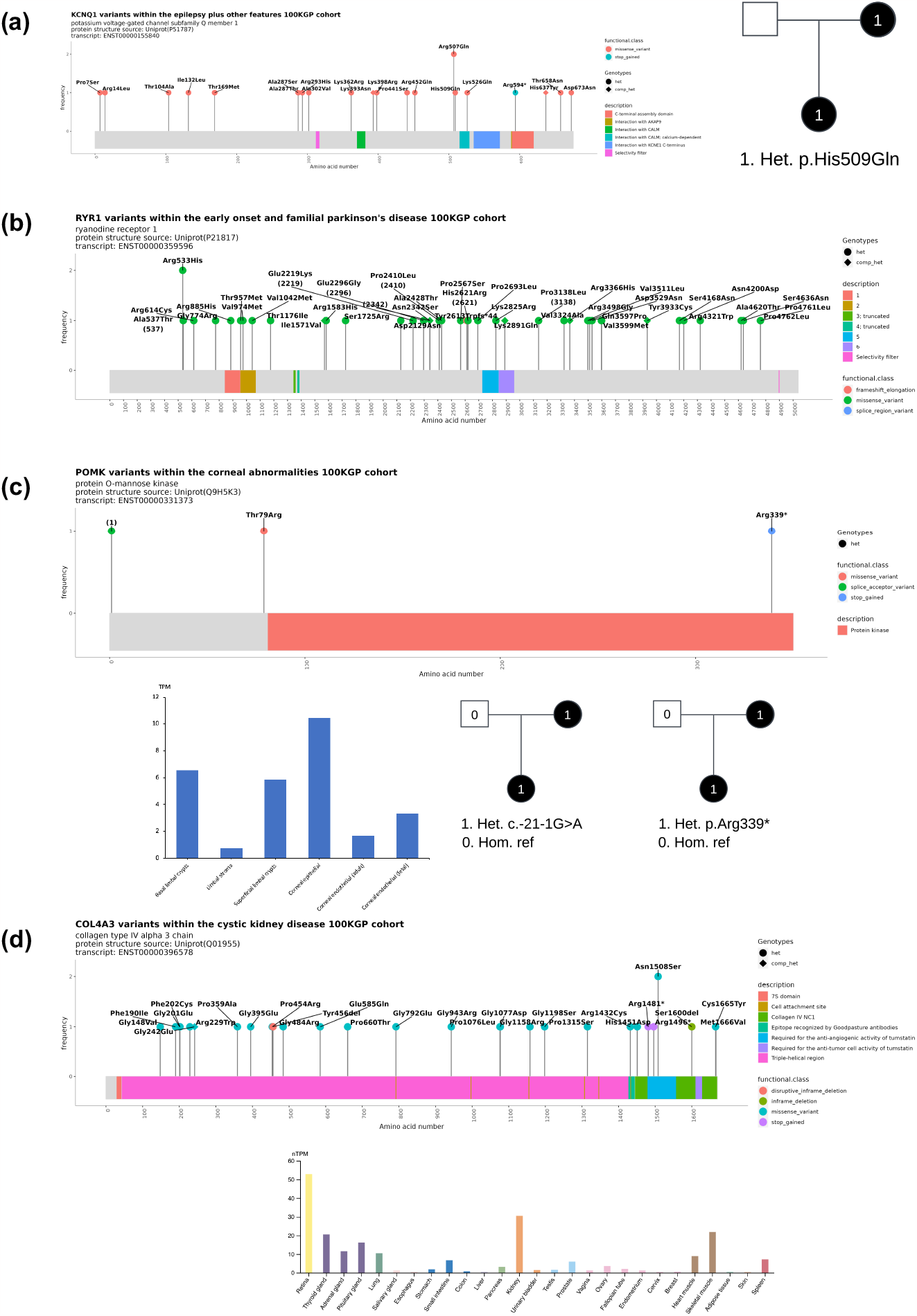
Evidence for associations based on location of rare, predicted pathogenic variants in the 100,000 Genomes Project cases, Genotype Tissue Expression (GTEx) Project or other RNA-seq relevant data, and co-segregation data. **(a)** epilepsy with *KCNQ1*, **(b)** early onset Parkinson’s disease with *RYR1*, **(c)** anterior segment ocular abnormalities with *POMK* **(d)** cystic kidney disease with *COL4A3*.

Furthermore, repeated ischaemic insults could explain cerebral abnormalities. Thus, our association may reflect, at least in part, detection of concealed long QT syndrome and may account for the aforementioned, distinct phenotype. Supporting this, two of the case variants (p.Ala302Val and p.Arg594*) are classified as known/likely pathogenic for long-QT syndrome in Clinvar. Nonetheless, another gene associated with long-QT syndrome, *KCNH2* has been more strongly associated with primary epilepsy ^35^ but was not detected in our analysis. Investigation of the cardiac phenotype in these cases of epilepsy will be required but these data are not available currently.

### Early onset Parkinson’s disease associated with *RYR1*

We identified a dominant association between variants in *RYR1* and ‘specific disease’ *early onset and familial Parkinson’s disease*. The association is driven by rare, predicted pathogenic variants (35 missense, 1 LoF) in 36 cases, located throughout the gene (**Fig. 4b**, OR = 2.1 [95% CI: 1.5 – 2.9], **Supplementary Table 2**). The gene is depleted for rare missense variants in gnomAD (o/e = 0.9 (0.87-0.93) and Z = 1.92). The 36 cases show strong phenotypic similarity to each other (mean PhenoDigm score from pairwise, reciprocal, non-self hits was 0.76). However, 407/1000 randomly sampled case sets of the same size achieved the same mean score or higher, suggesting the *RYR1* families do not have any phenotypic distinction from the other Parkinson’s disease (PD) cases based on the available HPO terms. *RYR1* functions as a calcium release channel and is associated with malignant hyperthermia, congenital myopathies and tremor pathophysiology ^36^. A mouse knockout shows various neurological phenotypes such as being unresponsive to tactile stimuli, abnormal posture, and paralysis ^37^. The GTEx substantia nigra gene co-expression network was examined for enrichment of 45 genes related to Mendelian forms of PD and complex Parkinsonism (green genes in PanelApp) as well as 151 genes linked to sporadic PD through Mendelian randomisation (Nalls et al., 2019). *RYR1* was contained within the substantia nigra ‘darkmagenta’ module. Although this module was not significantly enriched for PD-associated genes, it is interesting to note that the module contained multiple genes causally implicated in PD, including *PINK1* and *PARK2* amongst others (FDR adjusted p-value = 0.146). Furthermore, the module in which *RYR1* is located is highly enriched for gene ontology terms associated with mitochondrial function, a key process in PD pathophysiology (**Supplementary Fig. 1**).

### Anterior segment ocular abnormalities associated with *POMK*

We identified a dominant association between variants in *POMK* and ‘specific disease’ *corneal abnormalities*. The association is driven by rare, predicted pathogenic variants (2 LoF, 1 missense) in 3 cases with recorded phenotypes collectively suggestive of Anterior Segment Dysgenesis (ASD) (**Fig. 4c**, OR = 92.5 [95% CI: 26.8 – 319.6], **Supplementary Table 2**). ASD is a spectrum of developmental disorders affecting the anterior segment of the eye often with incomplete penetrance and/or variable expressivity ^38^. Co-segregation was apparent in two trios where a heterozygous, splice acceptor variant c.-21-1G>A (gnomAD v4.0.0 allele frequency (AF) = 0.000011) and a heterozygous, frameshift stop gain variant p.Arg339* (gnomAD v4.0.0 AF = 0.000001) are inherited from the affected mothers in the female probands (**Fig. 4c**). In the independent cohort described in **Supplementary Table 2**, one rare (gnomAD v4.0.0 AF = 0.000038), heterozygous missense variant p.Arg86His was identified in a mother and a son of a Czech family diagnosed with ASD, comprising 4 affected individuals across 3 generations. *POMK* is involved in the presentation of the laminin-binding O-linked carbohydrate chain of alpha-dystroglycan (a-DG), which forms transmembrane linkages between the extracellular matrix and the exoskeleton. Given the absence of corneal specific expression data in the GTEx Project, we interrogated publicly available bulk RNA-seq datasets ^39,40^ which showed expression across all corneal cell types analysed, with highest levels detected within the corneal epithelium (**Fig. 4c**). Bi-allelic (predicted LoF) mutations in *POMK* are associated with autosomal recessive muscular dystrophy-dystroglycanopathy (congenital with brain and eye anomalies), type A, 12 (OMIM:615249), a disease that also includes several ocular abnormalities (microphthalmia, buphthalmos, coloboma, retinal degeneration and cataract), suggesting *POMK* plays a crucial role in ocular development ^41^. Morpholino knockdown of the *pomk* gene in zebrafish has been reported to show multiple defects, including developmental ocular abnormalities ^41^. Whether the identified rare variants here could induce ASD via *POMK* haploinsufficiency, or by exerting dominant gain-of-function effects, deserves future investigation. With *POMK* apparently LoF-tolerant (pLI = 0) and ASD not been reported in carriers of muscular dystrophy-dystroglycanopathy type A, 12-associated variants, the latter seems more likely.

### Cystic kidney disease associated with *COL4A3*

We identified a dominant association between variants in *COL4A3* and ‘specific disease’ *cystic kidney disease* and another component of Type IV collagen, family member *COL4A1*, is already known to be associated with cystic kidney disease (Plaisier et al. 2007). The association was driven by rare, predicted pathogenic variants (27 missense, 2 LoF) in 29 cases, located throughout this kidney expressed gene (**Fig. 4d**, OR = 4.4 [95% CI: 2.9 – 6.6], **Supplementary Table 2**). *COL4A3* is constrained for rare missense variants in gnomAD (o/e = 0.77 (0.67-0.82) and Z = 1.83). The 29 cases show strong phenotypic similarity to each other (mean PhenoDigm score from pairwise, reciprocal, non-self-hits was 0.81) with cardiovascular phenotypes observed in some cases in addition to kidney cysts. 224/1000 randomly sampled case sets of the same size achieved the same mean score or higher, suggesting the *COL4A3* families do not form a phenotypically distinct group from the other cases based on HPO terms. Three of the cases involved duos where the variant was also observed in the other affected family member: (i) a ClinVar known pathogenic variant, p.Gly395Glu ^42^, in a brother and sister, a ClinVar VUS variant, p.Ser1600del, in a father and son, (iii) a ClinVar pathogenic variant, p.Arg1481*, in a mother and son. *COL4A3* is associated with dominant (van der Loop et al. 2000) and recessive (Mochizuki et al. 1994) forms of Alport syndrome with renal and hearing phenotypes, as well as a milder familial hematuria (Badenas et al. 2002). Kidney cysts in people with Alport syndrome/thin basement membrane nephropathy have previously been observed (Savige, Mack, et al. 2022). However, only one of our cases, with a p.Asn1508Ser variant reported with conflicting evidence in Clinvar, had any hearing impairment suggestive of classical Alport syndrome. Eleven of the cases have already been issued with molecular diagnoses in other cystic kidney genes (9 with *PKD1* and 2 with *PKD2*) within the 100KGP, raising the possibility of digenic inheritance or modification of incompletely penetrant variants. In the independent Irish cohort described in **Supplementary Table 2**, two related patients with chronic kidney disease are already diagnosed with rare, heterozygous *COL4A3* variants and two have *COL4A3* and missense *PKD1* VUS variants. A digenic form of Alport syndrome has been described with pathogenic *COL4A4*/*COL4A5* and milder *COL4A3* variants ^43^. This more severe, form of Alport syndrome is associated with increased incidence of proteinuria, hypertension and kidney failure is occasionally observed and the recommendation is to look for second variants in *COL4A3, COL4A4* and *COL4A5* in at-risk individuals (Savige, Lipska-Zietkiewicz, et al. 2022). In our cases with an existing cystic kidney disease molecular diagnosis, the renal phenotype is not as severe and pathogenic *COL4A4* and *COL4A5* variants are not observed suggesting the *COL4A3* variants are having a milder digenic effect or acting as modifiers with other cystic kidney genes.

## Discussion

In this study we have described a gene burden analysis of a large cohort of rare disease cases and identified 88 novel disease-gene associations after triaging of the statistically significant signals. Recent publications describe confirmation of five of these and we highlight a further seven with strong genetic and experimental evidence: hypertrophic cardiomyopathy associated with *DYSF* and *SLC4A3*; monogenic diabetes with *UNC13A*; epilepsy with *KCNQ1*; early onset Parkinson’s disease with RYR1; anterior segment ocular abnormalities with *POMK*; and cystic kidney disease with *COL4A3;* and. Further evidence is necessary before all the novel associations described here can be used clinically for diagnostics, counselling, and management. For example, addition of further functional study evidence would increase the score of 47/50 of the limited ClinGen classification evidence candidates shown in **Supplementary Table 2**, such that they would be re-classified as moderate. We are also pursuing the identification of variants in further independent cases through GeneMatcher to boost the ClinGen evidence category. Collection of additional affected family members for cases in collaboration with the original recruiting clinicians could also raise the category although this is likely to be more difficult, e.g., for a dominant association at least two large families with five affected members are needed to add one point of evidence.

Although our approach applied to a large, rare disease cohort has successfully highlighted numerous known associations and suggested many previously unreported associations, it is not without limitations. There is an assumption that variant calls are accurate and that cases are always recruited to the correct ‘specific disease’ category within the 100KGP, which is not always true as shown for *DCP1A* with heart disease and *HBB* with kidney stones respectively. Probands are assumed to be unrelated, but when we further investigated the association of *ADAMTS4* with hearing impairment (**Supplementary Table 2**), this was not the case with 3 of the 4 probands belonging to the same family. The *GATA2* signal was similarly diluted on further investigation with 4 of the 6 probands linked to the association being closely related (mother and 3 siblings). For maximal performance of a gene burden testing approach, there should be no pathogenic variants associated with the disease cases in the control samples but mis-recruitment or incomplete penetrance may mean this is not always true, otherwise we could have added this absolute condition prior to the statistical testing. Given the multi-disease nature of our analysis, we opted for a non-disease specific, less strict AF filtering strategy so as not to incur inflated false negatives and afterwards discarded, in our *in silico* triage, signals where > 33.3% of the contributing case variants were present in controls (179/316, 57%). Those many discarded signals could be the focus of a future clinical expert revised triage using a *disease-specific* AF filtering strategy such as the *maximum credible population AF* ^44^ where disease-specific prevalence, genetic and allelic heterogeneity, inheritance mode, penetrance, and sampling variance in reference datasets are considered to identify less arbitrary and/or unnecessarily lenient AF cutoffs.

The relatively small ORs for some of the associations in **Supplementary Table 2** suggest that some of the 88 associations may involve incomplete penetrance and that is known to be the case for kidney stones and *SLC34A3*. Incomplete penetrance will lead to the same/similar variants in controls and lower ORs and less significant signals, as well as possible loss of signal in multi-sample cases under Exomiser’s assumed full penetrance model. Finally, associations linked to gain-of-function variants may have been difficult to detect with our methodology, unless they clustered in the CCR regions we analysed.

A recent analysis of approximately the same cohort of rare disease patients using BeviMed found 241 known but only 19 novel associations ^24^. 115/190 of our known and only 5/88 of the novel signals were also detected by BeviMed: *UCHL1* associated with hereditary ataxia; *SLC34A3* and *HBB* associated with renal tract calcification; *TUFT1* associated with epidermolysis bullosa; *SRP9* with ductal plate malformations. The relatively small overlap in signals highlights a possible complementarity of the two methods to discover new disease-gene associations from the same cohort. As well as differences in the statistical approach, different case-control selection strategies were used.

There are 606 cases with no molecular diagnosis but with variant(s) contributing to one of the known disease-gene association signals, that had not already been considered and classified as VUS or benign in the diagnostic report, giving an upper bound on the increase in diagnostic yield from review of these variants of 1.7% (606 of 35,008 cases analysed). Furthermore, 456 molecularly unsolved cases had a variant contributing to one of the 88 novel associations and could potentially increase the diagnostic yield by 1.3% (456 of 35,008 cases analysed), if all genes were confirmed and the variants considered penetrant enough to be deemed pathogenic rather than just predictive. By making our analytical framework openly available for wider application to similar cohort data globally, we hope to substantially aid disease-gene discovery and new molecular diagnoses in rare Mendelian diseases in numerous other cohorts.

## Online Methods

### Rare disease genomes from the 100,000 Genomes Project

Patients with rare diseases and affected and unaffected family members were enrolled to the 100KGP through one of the 13 NHS Genomic Medicine Centres (GMCs) across England, Northern Ireland, Scotland and Wales ^8^. The recruiting clinicians assigned each proband to a specific disease (according to a hierarchical disease classification available within the project which is described below) and provided patient’s phenotypic data based on the HPO ^45^. An initial cohort of 74,061 genomes (35,548 single probands and larger families) from the rare disease pilot and main programme of the 100KGP (Data Release v.11) was available for analysis (March 2021). Genome sequencing was performed with the use of the TruSeq DNA polymerase-chain-reaction (PCR)–free sample preparation kit (Illumina) on a HiSeq 2500 sequencer, which generates a mean depth of 32× (range, 27 to 54) and a depth greater than 15× for at least 95% of the reference human genome. Whole-genome sequencing reads were aligned to the Genome Reference Consortium human genome build 37 (GRCh37) with the use of Isaac Genome Alignment Software. Family-based variant calling of single-nucleotide variants (SNVs) and insertion or deletions (indels) for chromosomes 1 to 22, the X chromosome, and the mitochondrial genome (mean coverage, 2814×; range, 142 to 16,581) was performed with the use of the Platypus variant caller ^46^. Quality control performed by Genomics England highlighted that 81 of the probands had been recruited and sequenced twice and these duplicates were removed from our cohort. In addition, the required data for our Exomiser-based gene burden analysis, e.g. recruited disease category and phenotypic terms, was not available for 16 families and these were also excluded from our cohort.

### Pool of rare, putative disease-causing variants for gene burden testing

The variant prioritisation tool Exomiser ^9^ (version 12.1.0 with default settings and latest 2007* (July2020) databases) was then run on all available 35,451 single proband and family-based variant call format (VCF) files to obtain a pool of rare, protein-coding, segregating and most predicted pathogenic (per each gene) variants to use in a rare-variant gene-based burden testing analysis for the discovery of novel rare Mendelian disease-gene associations as described below. Per each proband/family and each gene, Exomiser selected a single configuration of *contributing* variants, i.e. the most predicted (i.e. REVEL and MVP) pathogenic, rare (< 0.1% autosomal/X-linked dominant or homozygous recessive, < 2% autosomal/X-linked compound heterozygous recessive; using publicly available sequencing datasets including gnomAD) protein-coding homozygous/heterozygous variant or compound-heterozygote variants that segregated with disease for each possible mode of inheritance. Coding variants were selected by Exomiser by removing all those classified as FIVE_PRIME_UTR_EXON_VARIANT, FIVE_PRIME_UTR_INTRON_VARIANT,THREE_PRIME_UTR_EXON_VARIANT, THREE_PRIME_UTR_INTRON_VARIANT, NON_CODING_TRANSCRIPT_EXON_VARIANT,UPSTREAM_GENE_VARIANT, INTERGENIC_VARIANT,REGULATORY_REGION_VARIANT, CODING_TRANSCRIPT_INTRON_VARIANT, NON_CODING_TRANSCRIPT_INTRON_VARIANT, DOWNSTREAM_GENE_VARIANT. The Exomiser analysis did not return any candidate variants for 29 families, generally for larger families with multiple affected individuals where no rare, putative disease-causing variants remained after filtering, leading to an interim dataset size of 35,422 single probands and larger families and 5,733,899 Exomiser-based candidate variants (*Exomiser master dataset*). To control for false positive variant calls and/or relatively common variants within the project itself, we further discarded variants based on how often they were observed within the Exomiser master dataset itself (frequency > 2% for variants in a compound-heterozygote genotype, > 0.2% for mtDNA genome variants, > 0.1% for heterozygote/homozygote variants). This led us to discard data from 41 additional families. Finally, potentially digenic probands with more than one recruited disease category were discarded from the analysis, leading to a final analysis dataset of 35,008 families (40,584 probands and affected family members and 32,434 unaffected family members) and 4,676,866 Exomiser-based candidate heterozygote/homozygote variants and compound-heterozygote genotypes (**Supplementary Table 1 and Fig. 1**).

### Exomiser-based rare variant gene burden testing

A rare variant gene-based burden case-control analytical framework which exploits rare, putative disease-causing variants as annotated, filtered and scored by the variant prioritisation tool Exomiser has been used to identify novel rare Mendelian disease-gene associations. The framework has been described previously ^4^. Briefly, as to the application of the analytical framework to the rare disease component of the 100KGP, *cases* and *controls* were defined exploiting the hierarchical disease classification within the project itself where the recruiting clinicians assigned each proband to any of 226 ‘specific diseases’ (level 4); the ‘specific diseases’ are in turn grouped into less specific 91 ‘disease sub groups’ (level 3), each of which corresponds to one of 20 broad ‘disease groups’ (level 2) (**Supplementary Table 1**). A *case* set was then defined as all probands recruited under each of the 226 level 4 disease categories and its corresponding *control* set as all recruited probands except those under the level 2 category containing the specific level 4 disease, e.g. ‘hypertrophic cardiomyopathy’ cases were compared to all non-’cardiovascular disorders’ probands as controls. As to the gene burden testing, right-tailed Fisher’s exact tests were performed to assess the gene-based enrichment of variants in cases versus controls under four proband’s genotype scenarios (irrespective of the mode of inheritance): (i) presence of at least one rare, predicted LoF variant, (ii) presence of at least one rare, highly predicted pathogenic variant (Exomiser variant score >= 0.8, i.e. either LoF or missense variants predicted to be pathogenic) (iii) presence of at least one rare, highly predicted pathogenic variant in a constrained coding region (CCR) and (iv) presence of a rare, *de novo* variant (restricted to only trios or larger families where *de novo* calling was possible). To maintain statistical validity and power, the analysis was limited to those disease-gene associations where at least five cases exist for the specific disease tested and, per each of the four gene-based proband’s genotype scenarios above, where relevant variants in the gene were seen in at least four probands, of which at least one was a case. The Benjamini and Hochberg method ^47^ was used to correct for multiple testing; an overall false discovery rate (FDR) adjusted p-value (q-value) threshold of 0.05 was used for claiming statistically significant disease-gene associations to pursue for further triaging.

### Triaging

#### First in silico triage

The statistically significant associations were further filtered for those where (i) the gene was protein-coding as the Exomiser coding variant filtering settings also identified variants disrupting non-protein coding RNA genes, (ii) less than 33.3% of the variants driving the signal in the cases were present in the controls, (iii) for dominant, LoF signals there was gnomAD evidence for haploinsufficiency (gnomAD LOEUF < 0.5), and (iv) <= 50% of the cases driving the signal were already assigned a molecular diagnosis in other genes as part of the 100KGP routine diagnostic pipeline.

#### Second clinical expert (GeCIP) and in silico triage

An automated classification of the disease-gene associations according to ClinGen criteria(https://www.clinicalgenome.org/site/assets/files/9232/gene-disease_validity_standard_operating_procedures_version_10.pdf) was applied. The case-level variant score was calculated from scoring and summing all case variants that support a particular mode of inheritance for a disease-gene association. LoF variants (stop gain, frameshift or splice acceptor/donor) scored 1.5 points or 2 if *de novo* whilst others scored 0.1 points or 0.5 if *de novo*. A case-control study score of 5 points for an OR > 5, 4 points for OR > 3 or 3 points for OR < 3 was assigned. The larger of the case-level variant score or case-control study score was used as the genetic evidence score, capped at a maximum of 12 for those associations that had many supporting case variants. Experimental evidence categories were calculated using a variety of sources. Existing evidence for a gene function fitting the likely disease mechanism was assessed via PubMed searches using the disease and gene name and the background knowledge of the experts in the various disease-specific GeCIPs. Scores of 0, 1 or 2 were awarded depending on whether there was no, some tenuous or lots of evidence. Gene expression was assessed using GTEx Project data through the web portal of the Human Protein Atlas (https://www.proteinatlas.org/; ^48^ and/or publicly available relevant RNA-seq datasets ^39,40^, and a score of 0, 1 or 2 assigned for no, widespread or solely specific expression in the relevant disease tissue. Defaults of 1 point for protein-protein association evidence (high quality, direct experimental interactions (StringDB interactions with a score > 0.7 and experimental evidence) with genes on the disease panel from PanelApp ^14^ and 2 points for mouse/zebrafish evidence where there was some phenotypic similarity as calculated by Exomiser between the patient’s phenotypes and the mouse/zebrafish phenotypes where the orthologous gene was disrupted. The rounded sum of genetic and experimental evidence points was used to assign the final ClinGen classification of limited (0.1-6 points), moderate (7-11 points) or strong (12-18 points). Definitive evidence for an association is considered a score of 12-18 as well as convincing replication of the result in more than 2 publications over more than 3 years, and therefore not applicable here.

### Visual representation of variant location in lollipop plots

Visual representations of the variant locations within the protein were generated by extending the Mutplot software ^49^. The x-axis represents the amino acid chain and their annotated protein domain from UniProt. Each lolly indicates a variant by its protein change annotated on one single transcript (specified in the plot) and the frequency is shown on the y-axis. Its shape indicates the genotype found in the proband. The colour indicates the type of variant and the variant’s functional annotation. If the variant has both a p. change annotation and a number in parenthesis it means that the original p. change was annotated on a different transcript and the amino acid position in parenthesis indicates the re-annotation on the selected transcript. If the only annotation available indicates a number in parenthesis it means that the variant was in the non-coding region for that transcript, therefore the lolly was placed on the closest amino acid.

### PhenoDigm patient similarity comparisons

During assessment of some disease-gene associations, the phenotypic similarity between the probands driving the signal was calculated using their HPO term annotations and the Exomiser API to give a PhenoDigm ^50^ score between 0 and 1. The mean of the pairwise, reciprocal, non-self hits was calculated and compared to those obtained from 1000 iterations when the same number of probands was selected at random from the set of cases with that disease.

### Co-expression network analysis

Co-expression network analysis of our candidate genes and known genes linked to the potentially associated disease (green genes in PanelApp version 1.120) was performed using GTEx v6 tissue-specific modules and the CoExp tool accessible at https://rytenlab.com/coexp ^51^.

### Peripheral blood mononuclear cell (PBMC) expression analysis

RNA-seq data from PBMC cells collected from three volunteer donors was analysed (poly A-selected libraries, mean of two replicates untreated and two replicates treated with cycloheximide for 1hr to inhibit protein translation and mimic integrated stress response). In-house, R was used for DeSeq2 normalisations per library and calculation of the mean values for each transcript for the 2 replicate libraries per donor per condition. For global evaluations, across all 3 donors, the mean base value, log2fold change post cycloheximide and Benjamini-Hochberg adjusted p-value were then calculated.

### Gene and variant look up in independent rare disease cohorts

In a cohort of Irish renal patients (278 cystic kidney disease and 141 chronic kidney disease cases), rare (gnomAD MAF < 0.1%) LoF, missense, splicing or intronic variants were extracted for our novel renal disease-associated genes. A further cohort of over 3000 Dutch renal patients was queried for likely pathogenic/pathogenic variants in those genes using the Alissa bioinformatics pipeline. Similarly, a sequencing cohort of 212 participants with inherited corneal diseases, recruited in the UK and Czech Republic and pre-screened for known genetic causes, was interrogated for any rare variants in the candidate gene *POMK*.

## Data Availability

This research was made possible through access to data in the National Genomic Research Library, which is managed by Genomics England Limited (a wholly owned company of the Department of Health and Social Care). The National Genomic Research Library holds data provided by patients and collected by the NHS as part of their care and data collected as part of their participation in research. Secure access to data is possible by becoming a member of the Genomics England Research Network (formerly GECIP), a network of approved researchers with access to the Genomics England Research Environment. This secure workspace provides a place to carry out research on de-identified datasets in the National Genomics Research Library.

## Author contributions

M.C, A.E.D, P.L contributed to the analysis of the corneal patient cohort, evaluation of candidates and reviewing/editing manuscript. A.M.E, I.L contributed to the analysis of the Dutch cohort. S.E.F contributed to the analysis of the Exeter diabetes patient cohort, evaluation of candidates and reviewing/editing manuscript. P.C, E.E, O.T contributed to the analysis of the Irish cohort. K.A.B contributed to the analysis of the Irish cohort, evaluation of candidates and reviewing/editing manuscript. V.C, L.V, D.S developed the analysis pipeline, conducted analyses and cowrote the manuscript. M.C contributed to the development of analysis and reviewing/editing the manuscript. P.N.R contributed to the development of parts of the analysis pipeline. J.OB.J contributed to the development of parts of the analysis pipeline and reviewing/editing manuscript. H.R.G, H.H, S.L, R.N, A.T.P, N.R, S.K.W, S.L.Z contributed to the evaluation of candidates. E.F.M, G.A, E.R.B, D.B, M.R.B, K.B, L.F.C, P.C, S.J .D, M.F, D.P.G, R.S.H, S.A.H, H.M, H.M, A.D.M, W.G.N, E.A.OT, A.CM.O, S.R, O.SA, J.A.S, J.C.T, I.T, A.T, J.S.W, L.M.W contributed to the evaluation of candidates and reviewing/editing the manuscript. S.GR, A.R.H contributed to the expression analysis. M.R, C.L.S contributed to the expression analysis, evaluation of candidates and reviewing/editing manuscript. M.B, R.K, A.R.W, C.T contributed to the patient recruitment and phenotyping. C.GC, S.H contributed to reviewing/editing the manuscript.

## Acknowledgements

This research was made possible through access to data in the National Genomic Research Library, which is managed by Genomics England Limited (a wholly owned company of the Department of Health and Social Care). The National Genomic Research Library holds data provided by patients and collected by the NHS as part of their care and data collected as part of their participation in research. The National Genomic Research Library is funded by the National Institute for Health Research and NHS England. The Wellcome Trust, Cancer Research UK and the Medical Research Council have also funded research infrastructure. PL was supported by GACR 24-10324S. This research was funded in part by Aligning Science Across Parkinson’s [Grant numbers: ASAP-000478 and ASAP-000509] through the Michael J. Fox Foundation for Parkinson’s Research (MJFF). This is part of the NIHR Barts Biomedical Research Centre (Caulfield, Jones) portfolio of research. The analysis was supported by a grant from the NIH, National Institute of Child Health and Human Development 1R01HD103805-01 and PhD funding from the UCLH NIHR Hearing Health BRC, and we would like to dedicate this work to the wonderful supervision of the late Professor Maria Bitner-Glindzicz.

## Supplementary table/figure legends

**Supplementary Table 1. Demographics of the 100,000 Genomics Project cohort**.

**Supplementary Table 2. 88 novel, putative disease-gene associations discovered by gene burden testing of the 100,000 Genomes Project data**. Bolding in cells is used to indicate where there is co-segregation evidence from multiple families, experimental evidence (literature, mouse models or protein-protein interactions to related disease-genes), or the mode of inheritance (MOI) was mixed, recessive or gnomAD oe_lof < 0.5 for dominant, LoF signals or gnomAD oe_missense < 1 for dominant, score >= 0.8 signals.

**Supplementary Figure 1.**
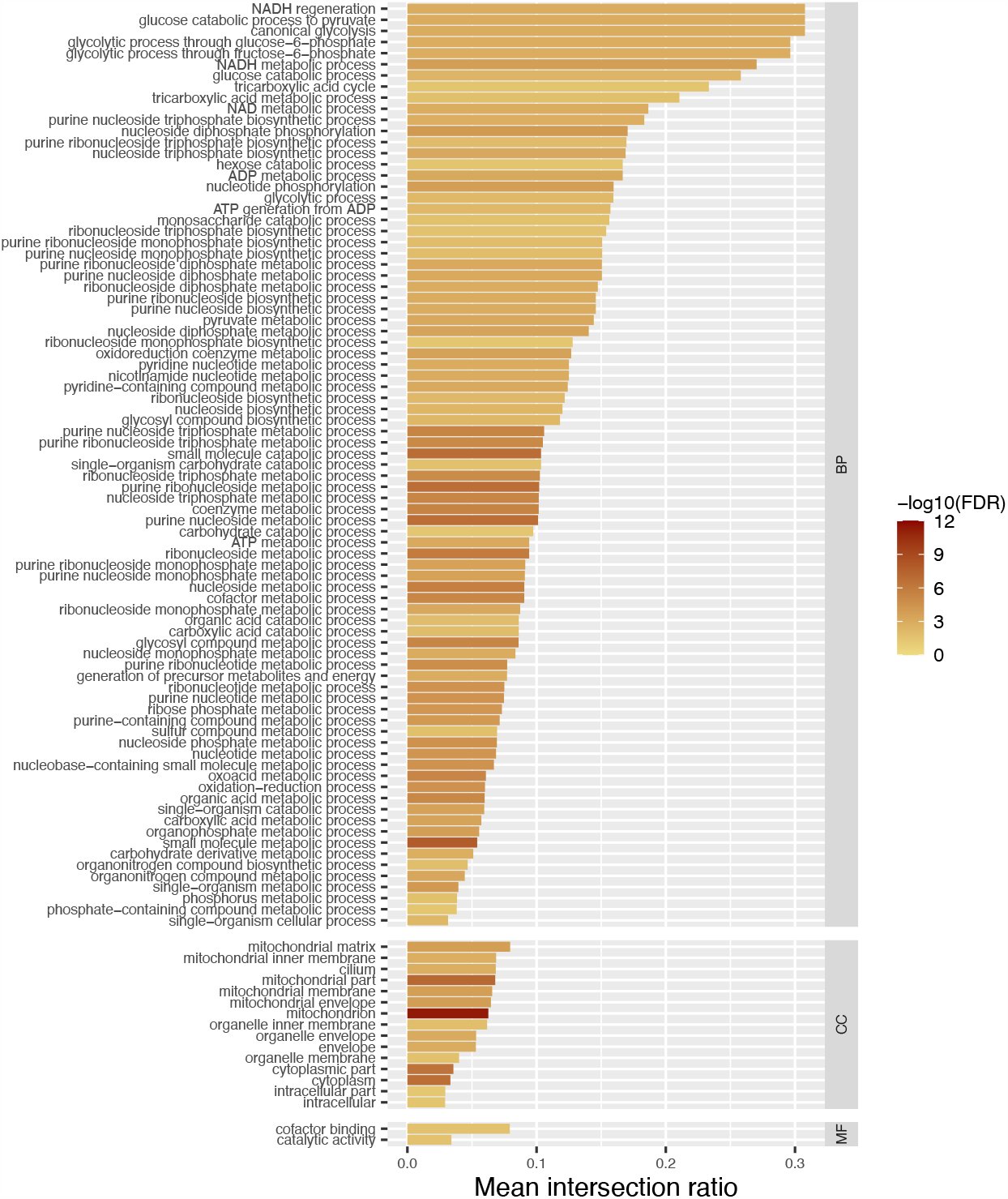
Gene ontology enrichment analysis of the GTEx substantia nigra gene co-expression network showing highly enrichment for terms associated with mitochondrial function, a key process in Parkinson’s disease pathophysiology.

